# Associations Between Lipid Traits and Breast Cancer Risk: A Mendelian Randomization Study in African Women

**DOI:** 10.1101/2024.09.23.24314169

**Authors:** Emmanuel Owusu Ansah, Foster Kyei, Caleb Frimpong Opoku, Andrews Danquah, Kwadwo Fosu, Emmanuel Boateng Agyenim, Daniel Sakyi Agyirifo

## Abstract

Blood lipids are associated with breast cancer. An increasing number of reports have attempted to explore the genetic connection between blood lipids and the risk of developing breast cancer. However, observational studies can be affected by confounding factors and reverse causation, which can compromise the reliability of the findings. We used univariate and multivariable two-sample mendelian randomization to explore the causal association between blood lipids and breast cancer. Summary-level data for lipid traits were obtained from the Africa Wits-INDEPTH partnership for Genomic Research (AWI-Gen) (N= 10,603, 58.5% of women). For breast cancer, we leveraged summary statistics from the most comprehensive Genome-wide Association Studies (GWAS) on breast cancer consisting of 18,034 cases and 22,104 controls of women of African ancestry. Our analysis suggests that genetically predicted triglycerides had a potential protective effect on breast carcinoma (OR = 0.73, 95% CI = 0.56, 0.95, FDR = 0.001). We found no evidence that genetically elevated levels of TC, HDL, and LDL may be associated with the risk of breast cancer TC (OR = 1.04; 95% CI, 0.93, 1.18; *FDR* = 0.029); HDL (OR = 1.29, 95% CI = 0.93, 1.79, FDR = 0.008); LDL (OR = 1.04, 95% CI = 0.90, 1.20, FDR = 0.036). Multivariate mendelian randomization analysis, which adjusted for the effects of TG, TC, LDL, and HDL, attenuated the observation of TG and breast cancer and also found no relationship between TC, HDL, LDL, and breast cancers. Furthermore, there was no evidence for a causal association between lipid traits and breast cancer subtypes. Our findings were robust in several sensitivity analyses. This study provides strong evidence that circulating TG may be associated with a decreased risk of breast cancer, while TC, LDL and HDL may not be related to the risk of breast cancer among African women.

## Introduction

Breast cancer (BC) is the second leading cause of cancer-related deaths among women worldwide, and understanding the causes of the factors is crucial to develop effective treatments **[1]**. Over the last 20 years, the annual increase in BC incidence has increased by approximately 0.33%, representing an increase from 876,990 cases to 2,002,350 **[2]**. Currently, Africa has the highest age-standardized breast cancer globally **[3–5]**. In particular, sub-Saharan African regions have the highest incidence rates **[2, 4, 6, 7]**. Given the best-studied risk factors for breast cancer, metabolic and behavioral risks remain the leading contributors to breast cancer-related deaths worldwide **[8–11]**. Furthermore, women with BRCA1 and BRCA2 mutations are significantly at increased risk of developing breast cancer, and these mutations have become a major focus of research **[12–15]**. Understanding and addressing these risk factors, especially in populations with higher breast cancer rates such as those of African descent, is essential. This knowledge is a crucial step toward creating targeted prevention strategies and improving treatment outcomes for women affected.

Cholesterol is a major metabolic factor associated with the risk of breast cancer, as elevated levels can contribute to tumor progression and metastasis **[16–19]**. Although growing experimental evidence suggests a possible link between plasma lipoprotein (the carriers of cholesterol) levels and breast cancer incidence **[17–19]**, it is not clear whether cholesterol directly influences breast cancer susceptibility in African populations. This knowledge gap is largely due to the limited number of epidemiological studies and genome-wide association studies (GWASs) conducted among women of African ancestry **[20]**. Observational studies have reported contrasting relationships— positive, negative, or null—between lipid levels and breast cancer risk **[21–23]**. For example, a comprehensive meta-analysis found that high-density lipoprotein (HDL) and low-density lipoprotein (LDL) can significantly increase Triple Negative and Luminal B BC, among African women, but they did not find an association with overall BC **[22]**. Another study reported an inverse association between breast cancer risk and increased levels of total cholesterol (TC), suggesting an interaction of circulating cholesterol levels with breast cancers **[24]**. However, these findings can be influenced by confounders that can obscure the relationship between cholesterol and breast cancer **[25, 26]**. Consequently, a high-powered causal inference analysis of lipids in BC is required. To our knowledge, there is no study that has systematically explored the potential genetic causation between serum lipid profiles and breast cancers in the African population.

Mendelian randomization (MR) analysis is a widely used technique to determine whether specific genetic variants have a causal effect on a particular trait or disease **[26, 28]**. MR leverages genetic variants that are randomly assigned at conception as instrumental variables (IV) **[29]**. By analyzing these genetic variants in relation to an exposure and its outcome using GWAS data, MR can identify whether the exposure directly causes the outcome. This approach minimizes the influence of confounders and biases, providing clearer insights into causal relationships **[28]**. In European populations, MR has been applied to establish causal links between various predictive factors and diseases, including lipid levels and risk of BC **[30, 31]**. For example, Orho-Melander and colleagues, using a small sample of 1,187 BC cases, found that increased HDL cholesterol and reduced triglycerides (TG) might increase the risk of breast cancer, while no association with LDL was observed **[31]**. On the contrary, another study that analyzed larger GWAS datasets reported nominal positive associations between LDL cholesterol levels and overall breast cancer risk, as well as between HDL cholesterol levels and estrogen receptor-positive breast cancer **[32]**. More recently, Johnson et al. also identified associations between HDL and LDL cholesterol and an increased risk of breast cancer but Jiang and colleagues found no statistically significant associations between any lipid traits and breast cancer **[33, 34]**.

Despite these promising findings, caution is warranted when generalizing these results to other populations, such as those of African ancestry, due to differences in genetic architecture between populations **[35]**. This highlights the need for further research to determine whether the causal relationships observed in European populations apply to other groups. In this study, we used MR to investigate whether genetically elevated lipid traits influence BC susceptibility, including both overall BC and specific subtypes, in African women. We leverage a recently published largest GWAS on breast cancer, which includes 18,034 cases and 22,104 controls from women of African ancestry, a dataset that has not yet been used in MR studies of breast cancer **[36]**. Briefly, we employed two-sample bidirectional MR to assess causal associations between circulating LDL, HDL, TC, and TG on the risk of total breast cancer, estrogen receptor-positive (ER-positive) breast cancer, estrogen receptor negative (ER negative) breast cancer, and triple-negative breast cancer (TNBC). We found possible risk-decreasing effects of increased TG that may have implications for overall breast cancer prevention.

## Methods

### Overview of the Study Design

The main analysis focused on the causal relationship of the lipid level with breast cancer and its molecular subtypes. Specifically, we investigated the causal effect of various blood lipids (TC, HDL, LDL, and TG), individually, on the development of breast cancer. These included 16 MR analyses using summary-level data from large-scale meta-analyses of genome-wide association studies. In addition, a reverse analysis was performed to understand whether breast cancer has a possible causal influence on lipid levels. Subsequently, a multivariate analysis was performed to establish the independent effect of each lipid type on the outcomes of breast cancer. The three main assumptions underlying MR, as illustrated in **Fig. 1**, are: first, instrumental variables (IVs) must be strongly associated with the exposure being studied; second, these IVs must not be linked to any confounding factors that could influence the relationship between the exposure and outcome; and third, genetic instruments should influence the outcome only through exposure. Details of sources of data used in this manuscript are summarized in **Table S1**. To reduce population stratification, all analyses were restricted to only African population.

**Figure 1.**
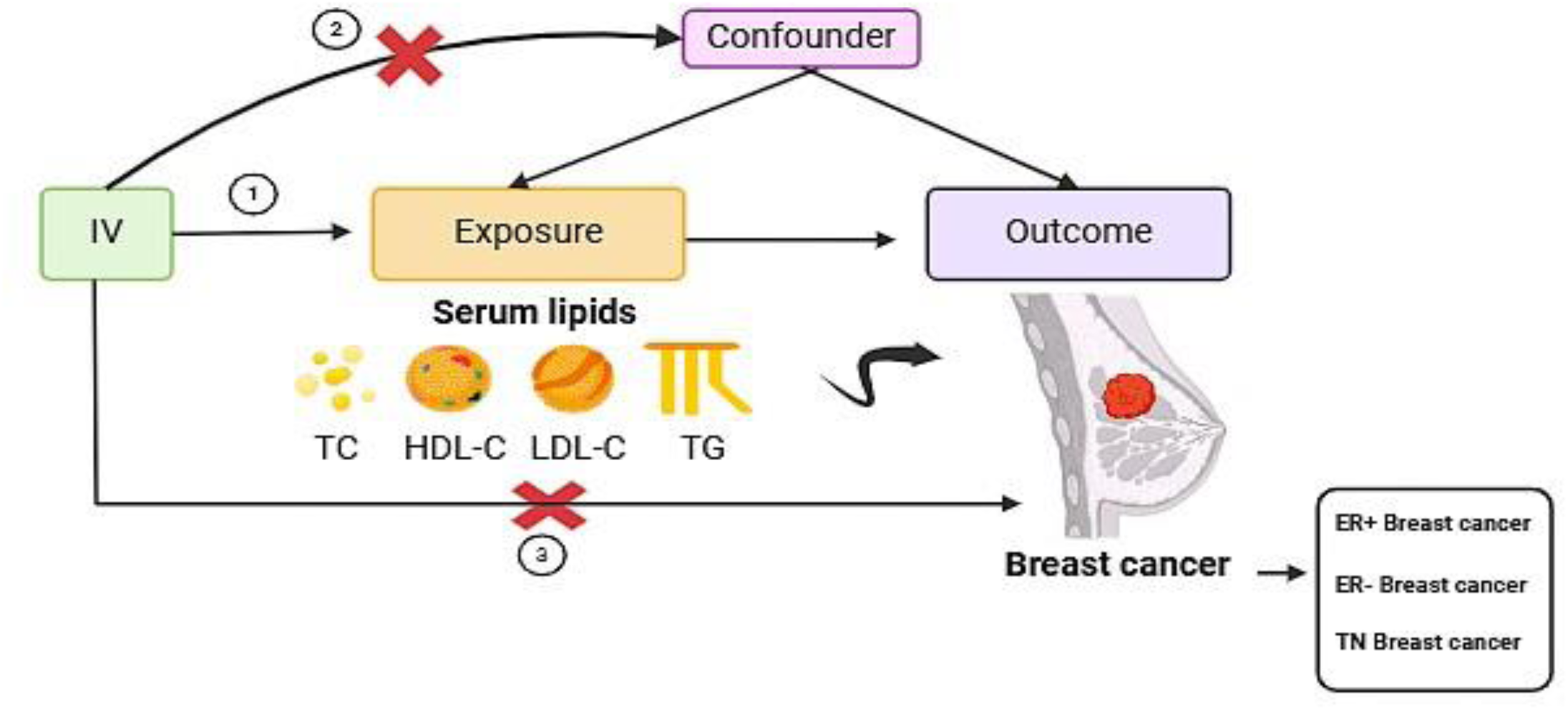
Study Overview. MR depends on three key assumptions. The exposures in the study were serum lipids, while the outcomes were breast cancer and its subtypes. TC total cholesterol, HDL-C high density lipoprotein cholesterol, LDL-C low density lipoprotein cholesterol, TG total triglyceride.

### GWAS data sources

Instrumental variables for this study were derived from large-scale genome-wide GWAS conducted on individuals of African ancestry. Genetic instruments for lipid traits were obtained from two key studies: the African Partnership for Chronic Disease Research (APCDR) and the Africa Wits-IN-DEPTH Partnership for Genomics Studies (AWI-Gen), which together included up to 24,215 participants. For breast cancer outcomes, we used a recent GWAS dataset comprising 18,034 cases and 22,104 controls. The outcomes examined included general breast cancer (18, 034), ER positive breast cancer (n = 9,304), estrogen ER negative breast cancer (n = 4,924) and TNBC (n = 2,860). These outcomes variables were defined by the African Ancestry Breast Cancer Genetic (AABCG) Consortium. Genotyping in these studies was performed using Illumina arrays or the Multi-Ethnic Genotyping Array (MEGA). Rigorous quality control (QC) procedures were applied to both datasets, including the removal of SNPs with missingness greater than 0.05, minor allele frequency (MAF) less than 0.01, and Hardy-Weinberg equilibrium (HWE) P-value less than 0.0001. Additional steps included imputation to the 1000 Genomes Project reference panel and adjustments for age, study design, and the first five principal genetic components. Ethical approval and participant consent were obtained in the original studies. All lipid data sets are available for download from the IEU OpenGWAS database (https://gwas.mrcieu.ac.uk/datasets/), and breast cancer datasets can be accessed from https://www.ebi.ac.uk/gwas/studies. Additional information on GWAS can be found in the original studies **[37, 38]**.

### Extraction of SNPs associated with lipid traits

We identified SNPs associated with each lipid trait from MRCIEU at a genome-wide significance threshold (p < 5 × 10^-8^). To ensure the independence of IVs, SNPs in the disequilibrium of the linkage (LD) with each other were removed using an LD pruning threshold of r^2^ = 0.001 and a kilobase (KB) threshold of 1000. The SNPs in the lipid datasets underwent screening, and consistency was ensured by harmonizing the direction of effect values between the exposure and outcome data. Ambiguous SNPs with incompatible alleles (e.g., A / G vs. A/C) were excluded from the analysis. Palindromic SNPs with intermediate allele frequencies (between 0.45 and 0.55) were also removed to minimize potential confounding effects that could violate the assumption of independence **[39]**. To assess the strength of selected SNPs, we calculated the F-statistic (F = beta^2^/se^2^) for each instrumental variable (IV). IVs with an F-statistic below 10 were considered weak instruments and therefore excluded from the analysis **[40]**.

### Quality Control and Data Standardization

Quality control of the breast cancer summary statistics was carried out following the guidelines outlined by Murphy et al. **[41]**. We utilized MungeSumstats, a Bioconductor R package, to standardize and process the summary statistics. MungeSumstats employs a series of automated quality control procedures to ensure data consistency and accuracy. Firstly, we standardized the column headers and checked the consistency of the data set, confirming that the alleles were correctly represented and aligned with the reference genome. Non-biallelic SNPs were removed, and any missing SNP IDs were imputed on the basis of base pair positions and chromosome numbers. Identified indels, as well as duplicated RSIDs and base pair positions, were excluded from the dataset. Next, we verified that the directionality of the effect alleles matched the reference genome. Any discrepancies in allele alignment were corrected by flipping the effect columns as needed. We also performed a change from hg38 to hg19 to align the summary statistics with the appropriate genomic coordinates, as the exposure dataset was stored in hg19. Further processing included renaming columns to accurately reflect minor or major allele frequencies and converting the summary statistics into the GenomicRanges format for better integration.

### Univariable Mendelian Randomization analysis

We conducted two-sample MR analyses using inverse variance weighted (IVW), weighted-median (WM), and MR-Egger models to estimate the casual relationship between lipid traits (TG, TC, HDL, and LDL) and the risk of breast cancers **[42–44]**. We use the random effect IVW method as the main effect size estimator **[45]**. The IVW regression model combines genetic variant-specific causal estimates weighted by the inverse of their variances **[42]**. The approach assumes the validity of the genetic instruments under the assumptions of no heterogeneity and no horizontal pleiotropy and, therefore, provides robust estimates of the causal effects. In the case of heterogeneity but no horizontal pleiotropy, the weighted median method was applied **[44]**. This method provides reliable estimates of the causal effect if at least half the weight comes from valid variants. However, when heterogeneity, or variation in causal estimates across genetic variants, and horizontal pleiotropy, where genetic variants influence multiple traits, were detected in our MR analyses, we performed the MR-Egger regression method for our analysis. Specifically, MR-Egger regression allows for an intercept term, which estimates the average pleiotropic effect across all variants used as instruments, to address biases induced by pleiotropy. Other MR algorithms, such as those for the simple median and simple mode described by Bowden, were also applied in this study to further assess the robustness of these findings.

### Multivariable Mendelian Randomization analysis

Since lipid traits are genetically related, we use MVMR to assess the direct effects of lipid traits on breast cancer outcomes, following the method described by Sanderson et al. **[46, 47]**. Here, we retrieved genetic-associated variants for all the exposures across their summary datasets. The SNPs for all traits (TC, HDL, LDL and TG) were combined and we then filtered for genome-wide significance (P < 5 × 10^−8^) and for linkage disequilibrium (r^2^ < 0.001). To test for the presence of weak instruments, we evaluated the strength and validity of IVs. We also performed horizontal pleiotropy testing using conventional Q-statistic estimation.

### Sensitivity Analyses

Heterogeneity among genetic variants was assessed using Cochrane’s Q value of the IVW method, with p < 0.05 indicating significant heterogeneity **[48]**. We used the MR-Egger intercept to check whether horizontal pleiotropy influenced our results **[49]**. A p-value greater than 0.05 suggested that pleiotropy was not a significant factor. Furthermore, MR-PRESSO was used to detect and correct outliers in the analysis **[50]**. The MR-PRESSO approach is derived from the IVW method but includes the removal of genetic variants whose specific causal estimates deviate from those of other variants **[50]**. Nonetheless, a leave-one-out approach was employed to evaluate the effect of each exposure SNP on the outcome of the MR analysis **[51]**. We achieved this by removing variant one by one from the analysis and re-estimating the causal effect. The results were presented as odds ratios (OR) and 95% confidence intervals (CI), providing an estimate of how lipid traits influence the probability of developing breast cancer **[52]**. P-value < 0.05 suggests that the observed association between lipid trait (risk factor) and the likelihood of developing breast cancer (outcome) is unlikely to be due to chance alone. To account for multiple tests in our analyses, we applied the false discovery rate (FDR) based on the Benjamini and Hochberg method (P value < 0.05/16 = 0.003125) **[53]**. All statistical analyses were performed in the R software version 4.2.2, using packages including TwoSampleMR, MVMR, MR-PRESSO and MendelianRandomization **[54–56]**.

## Results

### Single-trait MR in BC

In our study, we investigated the relationship between four lipid traits (i.e., TC, HDL-cholesterol, LDL-cholesterol and TGs), and breast cancer risk using data from the AWI-GEN study. In the forward analysis, we identified 16 SNPs, 8 SNPs, 14 SNPs and 6 SNPs associated with TC, HDL, LDL, and TG, respectively, at a genome-wide significance level (P < 5 × 10−8) (**Table 1**). In reverse analysis, 6 SNPs related to BC were selected as IVs. F statistics for all selected traits exceeded 10, indicating that there was no evidence of weak instrument bias. These SNPs explained approximately 0.2, 0.4, 3.6 and 3.7% of the variance of TG, HDL, TC and LDL, respectively **(Table 1)**. Information on SNPs for the exposure and outcome is listed in **Table S1**.

**Table 1.** Characteristics of the exposure variables. nSNPs, number of SNPs.

| Type | GWAS ID | Trait | nSNPs | F-statistics | R<sup>2</sup> (%) | Sample size | Population | PMID | Year |
| --- | --- | --- | --- | --- | --- | --- | --- | --- | --- |
| Exposure | ebi-a-GCST90101747 | Total Cholesterol | 16 | 78.8 | 3.6 | 24612 | African, females, males | 35546142 | 2022 |
|  | ebi-a-GCST90101746 | High Density Lipoprotein | 8 | 35.7 | 0.4 | 24616 | African, females, males | 35546142 | 2022 |
|  | ebi-a-GCST90101745 | Low Density Lipoprotein | 14 | 169.5 | 3.7 | 24515 | African, females, males | 35546142 | 2022 |
|  | ebi-a-GCST90101748 | Triglyceride | 6 | 47.3 | 0.2 | 24600 | African, females, males | 35546142 | 2022 |
|  | GCST90296719 | Breast cancer | 6 | 2073 | 0.8 | 18034 | African, females | 38741014 | 2024 |

In univariable MR analyses, triglyceride was found to be a protective factor against breast cancer. The IVW method indicated that 1- standard-deviation increase in TG was associated with an approximately 0.7-fold decrease in the risk of overall breast cancer (OR = 0.73, 95% CI = 0.56–0.95, FDR = 0.001; **Fig 2**, **Table 2**). To further confirm our results, we conducted additional MR analyses using various methods, including MR-PRESSO, IVW fixed effect method, MR-Egger, weighted median, and robust adjusted profile scores (RAPs) methods. The negative association was consistent across all these methods **(Fig. 3 and S2)**.

**Figure 2.**
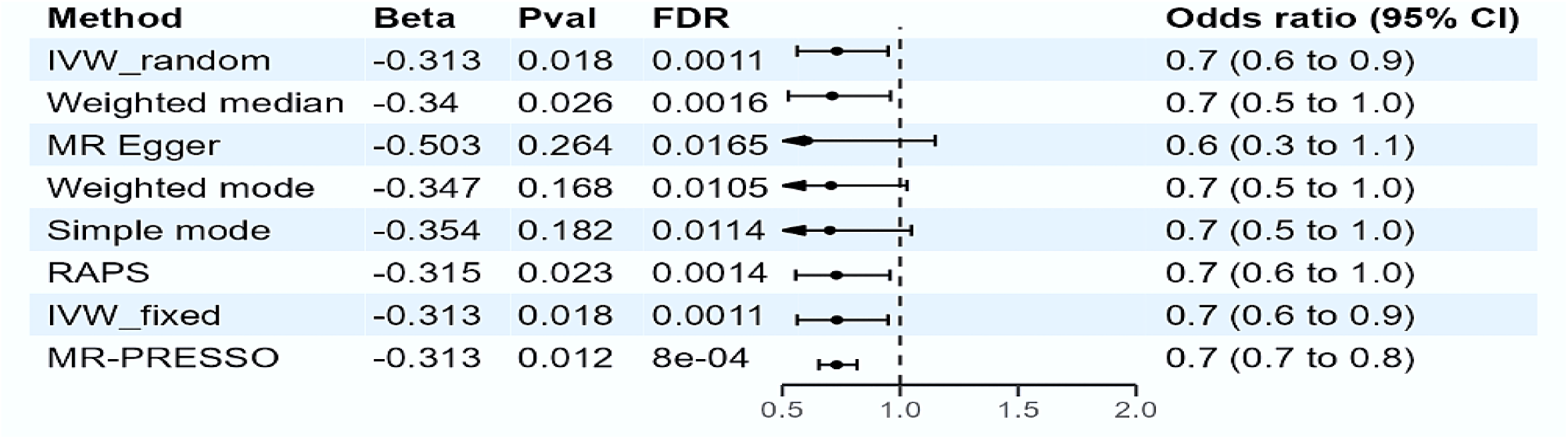
Estimates of triglyceride levels on the risk of breast cancer. Note: FDR, False Discovery Rate; Pval, P-value, CI, Confidence interval; IVW_random, random effect inverse-variance weighted; IVW_fixed, fixed effects inverse-variance weighted; MR-PRESSO, MR-pleiotropy residual sum and outlier; RAPS, robust adjusted profile score.

**Figure 3.**
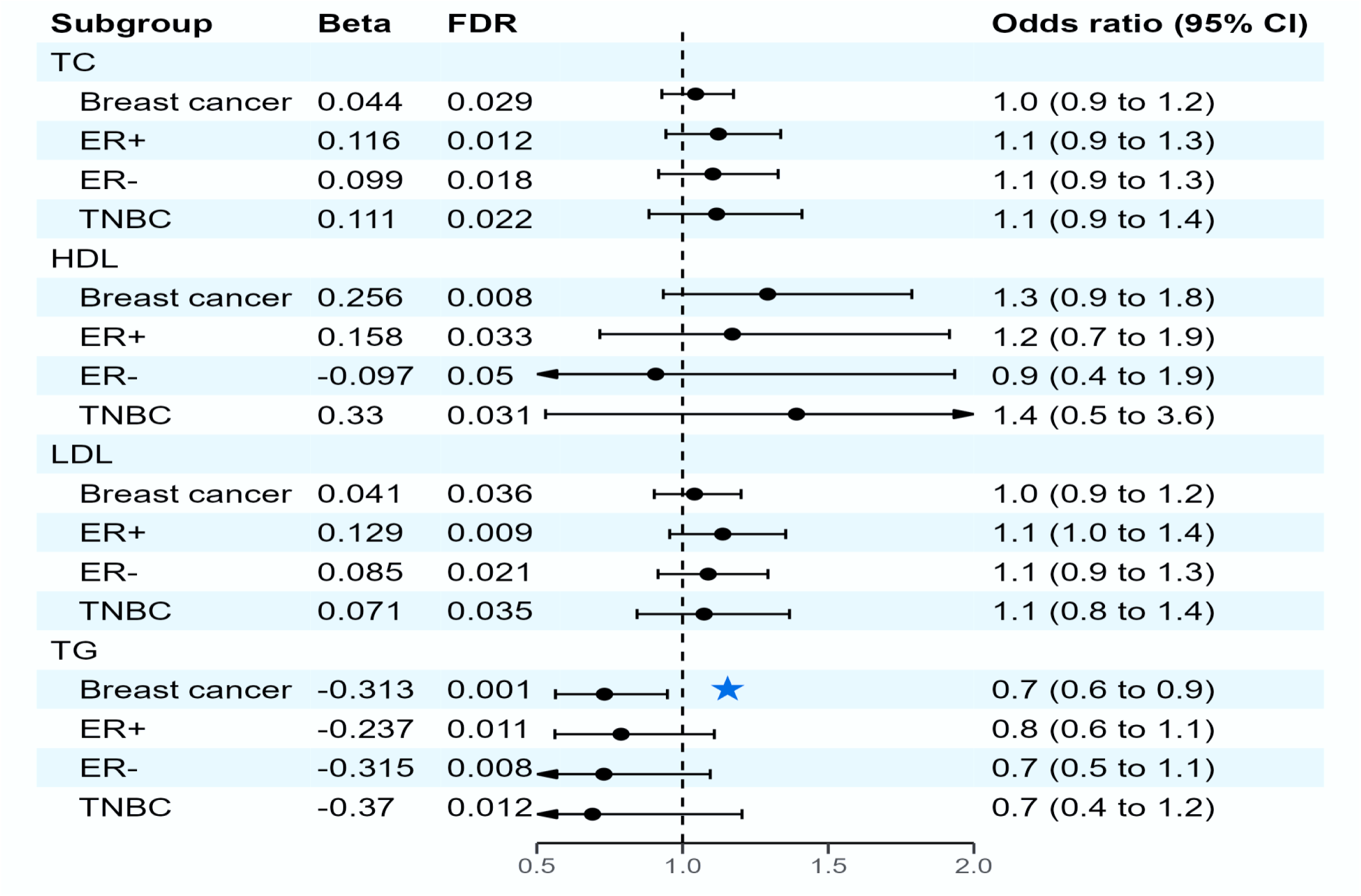
Estimates of triglyceride levels on the risk of breast cancer and its subtypes. Note: FDR, False Discovery Rate; CI, Confidence interval; TC, Total Cholesterol; HDL, High Density Lipoprotein; LDL, Low Density Lipoprotein; TG, Triglycerides; ER+, Estrogen Positive; ER-, Estrogen Negative; TNBC, Triple Negative breast cancer. Colored star indicates significant association

**Table 2.**
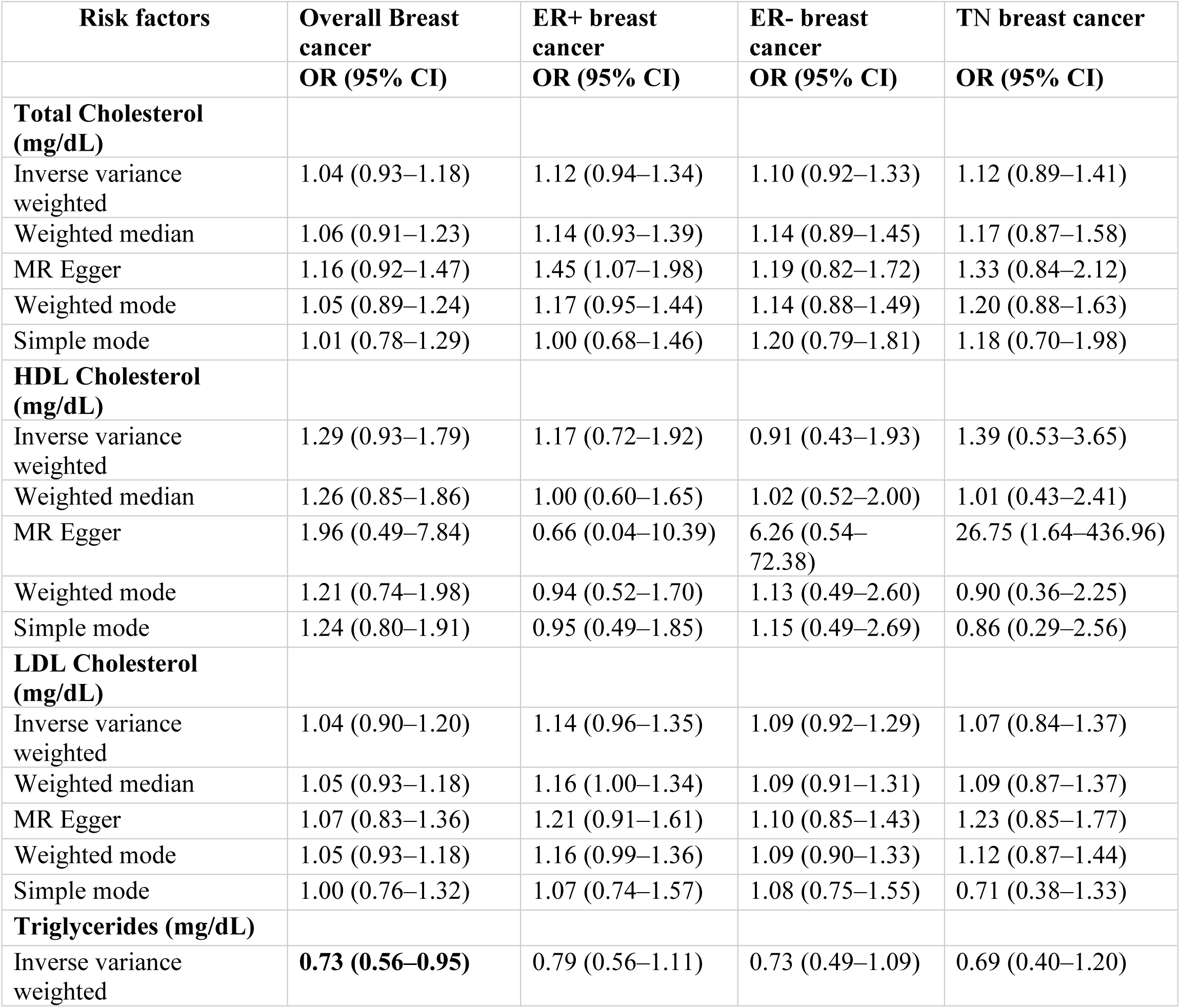
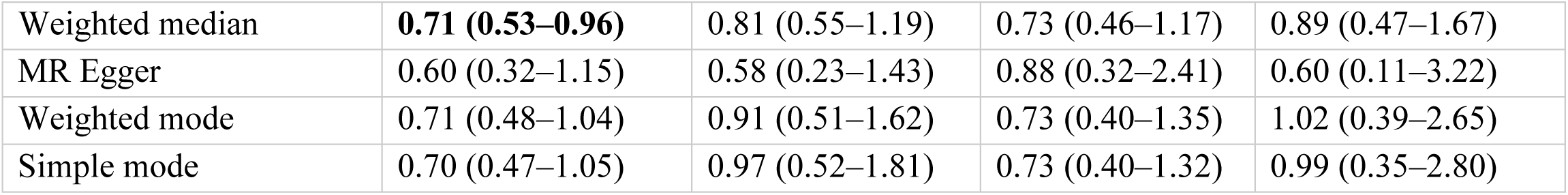
MR results for the relationship between Lipid traits and breast cancers. The MR analysis was performed through the TwoSampleMR packages (version 0.6.6) in R (version 4.2.2). All statistical tests were two-sided. P < 0.05 was considered significant. OR, odds ratio; CI, confidence interval; ER+, Estrogen Positive; ER-, Estrogen Negative; TN, Triple Negative.

| **Risk factors** | **Overall Breast cancer** | **ER+ breast cancer** | **ER- breast cancer** | **TN breast cancer** |
| --- | --- | --- | --- | --- |
|  | **OR (95% CI)** | **OR (95% CI)** | **OR (95% CI)** | **OR (95% CI)** |
| **Total Cholesterol (mg/dL)** |  |  |  |  |
| Inverse variance weighted | 1.04 (0.93–1.18) | 1.12 (0.94–1.34) | 1.10 (0.92–1.33) | 1.12 (0.89–1.41) |
| Weighted median | 1.06 (0.91–1.23) | 1.14 (0.93–1.39) | 1.14 (0.89–1.45) | 1.17 (0.87–1.58) |
| MR Egger | 1.16 (0.92–1.47) | 1.45 (1.07–1.98) | 1.19 (0.82–1.72) | 1.33 (0.84–2.12) |
| Weighted mode | 1.05 (0.89–1.24) | 1.17 (0.95–1.44) | 1.14 (0.88–1.49) | 1.20 (0.88–1.63) |
| Simple mode | 1.01 (0.78–1.29) | 1.00 (0.68–1.46) | 1.20 (0.79–1.81) | 1.18 (0.70–1.98) |
| **HDL Cholesterol (mg/dL)** |  |  |  |  |
| Inverse variance weighted | 1.29 (0.93–1.79) | 1.17 (0.72–1.92) | 0.91 (0.43–1.93) | 1.39 (0.53–3.65) |
| Weighted median | 1.26 (0.85–1.86) | 1.00 (0.60–1.65) | 1.02 (0.52–2.00) | 1.01 (0.43–2.41) |
| MR Egger | 1.96 (0.49–7.84) | 0.66 (0.04–10.39) | 6.26 (0.54–72.38) | 26.75 (1.64–436.96) |
| Weighted mode | 1.21 (0.74–1.98) | 0.94 (0.52–1.70) | 1.13 (0.49–2.60) | 0.90 (0.36–2.25) |
| Simple mode | 1.24 (0.80–1.91) | 0.95 (0.49–1.85) | 1.15 (0.49–2.69) | 0.86 (0.29–2.56) |
| **LDL Cholesterol (mg/dL)** |  |  |  |  |
| Inverse variance weighted | 1.04 (0.90–1.20) | 1.14 (0.96–1.35) | 1.09 (0.92–1.29) | 1.07 (0.84–1.37) |
| Weighted median | 1.05 (0.93–1.18) | 1.16 (1.00–1.34) | 1.09 (0.91–1.31) | 1.09 (0.87–1.37) |
| MR Egger | 1.07 (0.83–1.36) | 1.21 (0.91–1.61) | 1.10 (0.85–1.43) | 1.23 (0.85–1.77) |
| Weighted mode | 1.05 (0.93–1.18) | 1.16 (0.99–1.36) | 1.09 (0.90–1.33) | 1.12 (0.87–1.44) |
| Simple mode | 1.00 (0.76–1.32) | 1.07 (0.74–1.57) | 1.08 (0.75–1.55) | 0.71 (0.38–1.33) |
| **Triglycerides (mg/dL)** |  |  |  |  |
| Inverse variance weighted | **0.73 (0.56–0.95)** | 0.79 (0.56–1.11) | 0.73 (0.49–1.09) | 0.69 (0.40–1.20) |
| Weighted median | **0.71 (0.53–0.96)** | 0.81 (0.55–1.19) | 0.73 (0.46–1.17) | 0.89 (0.47–1.67) |
| MR Egger | 0.60 (0.32–1.15) | 0.58 (0.23–1.43) | 0.88 (0.32–2.41) | 0.60 (0.11–3.22) |
| Weighted mode | 0.71 (0.48–1.04) | 0.91 (0.51–1.62) | 0.73 (0.40–1.35) | 1.02 (0.39–2.65) |
| Simple mode | 0.70 (0.47–1.05) | 0.97 (0.52–1.81) | 0.73 (0.40–1.32) | 0.99 (0.35–2.80) |

There was no evidence that TC was associated with the odds of overall breast cancer (OR = 1.04; 95% CI, 0.93–1.18; *FDR* = 0.029) **(Table S2)**. Similarly, we did not observe any relationship with HDL or LDL-cholesterol, and the risk of breast cancer **(Table S2**). The effect directions from the other five methods were consistent with IVW **(Table S2**). Estimates of all causal associations between lipids and overall breast cancer are shown in Figure **2**. In a reciprocal single-trait MR test, we did not observe a relationship between the genetically determined risk of BC in each lipid trait **(Table S12)**.

The sensitivity analysis did not indicate significant heterogeneity between the SNPs, as shown by the Cochran Q test **(Table 3)**. The MR Egger intercept P-values were all above 0.05, showing no evidence of horizontal pleiotropy **(Table 3)**. Additionally, our leave-one-out analysis revealed that no single SNP significantly influenced overall casual estimates **(Figs. S5-S8, Table S5-S8).** Scatter plots for MR analyses are presented in **Figs. S1–S4**. Finally, the results of the MR-PRESSO analysis did not show outliers **(Table 3)**.

**Table 3.** MR results on heterogeneity and horizontal pleiotropy. Note. Q Cochran Q statistics; SE Standard Error; P P-value; TC, Total Cholesterol; HDL, High Density Lipoprotein; LDL, Low Density Lipoprotein; TG, Triglycerides

|  |  | *Heterogeneity test* |  |  |  |  |  | *Pleiotropy test* |  |  | *MRPRESSO* |
| --- | --- | --- | --- | --- | --- | --- | --- | --- | --- | --- | --- |
|  |  | IVW |  |  | MR-Egger |  |  | MR-Egger Intercept |  |  | global test |
| exposure | outcomes | *Q* | *Q_df* | *Q_pval* | *Q* | *Q_df* | *Q_pval* | *intercept* | *SE* | *P* | *P* |
| TC | Overall BC | 8.793 | 9 | 0.457 | 7.762 | 8 | 7.762 | -0.013 | 0.013 | 0.340 | 0.540 |
| HDL | Overall BC | 0.494 | 2 | 0.781 | 0.123 | 1 | 0.726 | -0.028 | 0.046 | 0.652 | - |
| LDL | Overall BC | 6.803 | 4 | 0.147 | 6.664 | 3 | 0.083 | -0.005 | 0.020 | 0.818 | 0.479 |
| TG | Overall BC | 0.542 | 3 | 0.910 | 0.138 | 2 | 0.933 | 0.019 | 0.029 | 0.590 | 0.935 |
| TC | ER+ BC | 12.452 | 9 | 0.189 | 8.616 | 8 | 0.376 | -0.032 | 0.017 | 0.096 | 0.220 |
| HDL | ER+ BC | 2.959 | 2 | 0.228 | 2.517 | 1 | 0.113 | 0.039 | 0.092 | 0.747 | 0.220 |
| LDL | ER+ BC | 6.324 | 4 | 0.176 | 5.686 | 3 | 0.128 | -0.013 | 0.023 | 0.602 | 0.364 |
| TG | ER+ BC | 3.281 | 3 | 0.350 | 2.572 | 2 | 0.276 | 0.031 | 0.041 | 0.535 | 0.439 |
| TC | ER- BC | 3.930 | 9 | 0.916 | 3.724 | 8 | 0.881 | -0.009 | 0.020 | 0.662 | 0.924 |
| HDL | ER- BC | 4.455 | 2 | 0.108 | 1.264 | 1 | 0.261 | -0.130 | 0.082 | 0.358 | - |
| LDL | ER- BC | 1.388 | 4 | 0.846 | 1.365 | 3 | 0.714 | -0.003 | 0.021 | 0.889 | 0.919 |
| TG | ER- BC | 0.563 | 3 | 0.905 | 0.403 | 2 | 0.817 | -0.018 | 0.046 | 0.728 | 0.918 |
| TC | TNBC | 3.568 | 3 | 0.312 | 3.514 | 2 | 0.173 | -0.028 | 0.030 | 0.409 | 0.634 |
| HDL | TNBC | 4.553 | 2 | 0.103 | 0.011 | 1 | 0.917 | -0.198 | 0.093 | 0.279 | - |
| LDL | TNBC | 4.957 | 4 | 0.292 | 3.798 | 3 | 0.284 | -0.028 | 0.030 | 0.409 | 0.525 |
| TG | TNBC | 3.568 | 3 | 0.312 | 3.514 | 2 | 0.173 | -0.018 | 0.046 | 0.728 | 0.349 |

### MR with outcome stratified by ER status

We performed MR analysis to test the relationship between genetically influenced lipids and BC risk stratified by ER-positive, ER- negative and TNBC status. In these stratified analyses, we found no association between TC, HDL, LDL, and TG and all the three subtypes of BC **(Table S3-S5)**. A heterogeneity test found no evidence to reject the null hypothesis of homogeneity between cancer subtypes (e.g., HDL: Cochran Q = 2.959, P = 0.228; **Table 3**). Therefore, we did not observe any substantive differences in the relationship of any lipid trait to ER+, ER− or TNBC.

### Multivariable MR

To test whether the association between lipid traits and breast cancer risk remains robust in the presence of other lipid factors (total cholesterol, high-density lipoprotein and low-density lipoprotein), we performed a MVMR analysis. In this approach, we used the four lipid traits as exposures and assessed their independent impact on the risk of breast cancer. We compared the IVs used for the lipid traits and identified eight overlapping IVs. Of these, 7 were shared between TC and LDL, while 1 IV overlapped between TG and HDL **(Fig. 4)**. We excluded overlapping SNPs, retaining 9, 7, 7, and 5 unique IVs for TC, LDL, HDL, and TG, respectively **(Fig. 4)**. These unique IVs were then used in the MVMR analysis to assess the independent effects of each lipid trait on the risk of BC. We did not observe significant relationships between genetically influenced HDL, LDL, TC, and TG with BC (HDL: OR = 0.113, 95% CI = 0.001-14.950, P = 0.415; LDL: OR = 3.048, 95% CI = 0.396-23.361, P = 3.048; TC: OR = 1.716, 95% CI = 0.416–7.059, P = 0.483; TG: OR = 6.359, 95% CI = 0.050-81.530, P = 0.398) (**Table S10**). Additionally, no association was detected when we considered the molecular subtypes (**Table S10**). A test for heterogeneity revealed no significant among the IVs (P > 0.05) **(Table S11).** Interestingly, our results were consistent with the univariable MR except for TG whose negative causal association with overall BC was abrogated after accounting for the interrelationships between these lipid traits (**Table S10**).

**Figure 4.**
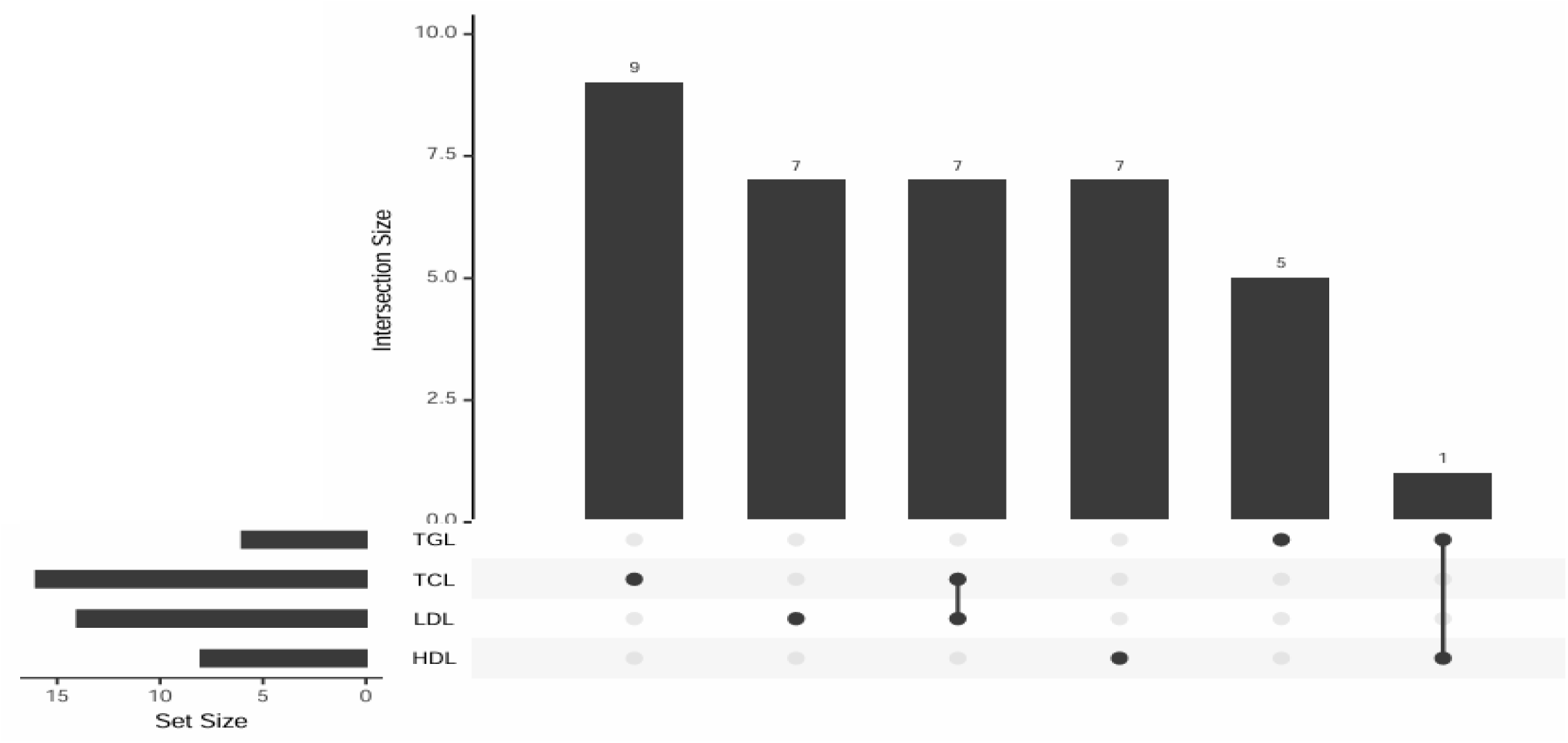
Overlap of instrumental variables between lipid traits. TC, Total Cholesterol; HDL, High Density Lipoprotein; LDL, Low Density Lipoprotein; TG, Triglycerides

## Discussion

To our knowledge, this is the first study using MR techniques to examine the causal relationship between plasma lipid levels and breast cancer risk among African populations, including molecular subtypes—ER+, ER- and TNBC. In the present study, we found that genetically elevated triglyceride levels were associated with a reduced risk of overall breast cancer. Our results are in line with other scientific reports **[32–34, 57–61]**. In experimental research, there is a claim that triglycerides could protect against breast cancer by shielding cells from fatty acid damage **[62]**. This protective effect was seen in other types of cancer, such as clear cell renal cell carcinoma (ccRCC) **[63]**. However, it is important to note that this finding contrasts with some other research. For instance, studies that included menopausal status in the analysis discovered an increase in TG levels which in turn elevated breast cancer risk **[64–67]**. This striking difference indicates that menopausal status could markedly influence the relationship between triglycerides and the risk of breast cancer. Another observation was that the association between TG and BC was attenuated in the multivariate analysis. These changes possibly mean that TG levels might not independently affect breast cancer risk **[60]**. Additionally, we also found no significant associations between total cholesterol, LDL, or HDL and breast cancer risk. Even when we stratified by molecular subtypes, the results remained unchanged. These results were consistent in various sensitivity analyses.

Generally, research on the association between serum cholesterol levels and breast cancer risk has produced conflicting results. Although some studies suggest positive connection **[68, 69]**, others provide an inverse relationship **[24, 58, 70]**. But our study did not find any significant changes in total cholesterol levels among breast cancer cases. Several studies have not reported no significant link between cholesterol levels and breast cancer risk **[59, 71–73]**. However, these inconsistencies in research outcomes can be attributed to various factors such as lifestyle, age, and racial disparity of the study participants **[74–76]**. Another explanation could be the metabolic status of patients, as conditions such as diabetes mellitus, insulin resistance, and obesity can influence cholesterol profiles and contribute to the development and progression **[77–79]**.

Cholesterol is transported in the body by LDL and HDL, and dysregulated levels of these lipoproteins have been implicated in breast cancer **[80–82]**. Studies using cell lines and animal models have shown that LDL can promote breast cancer cell survival, proliferation, and migration **[83, 84]**. In particular, Gallagher and colleagues demonstrated that elevated circulating LDL contributes to the growth of triple-negative and HER2-overexpressing breast cancers **[85]**. However, our MR study, along with several observational studies, did not find an association between LDL and breast cancer risk **[86–88]**. Interestingly, one study found that genetically higher LDL-C was associated with an increased risk of breast cancer **[32, 33]**, but other researchers using similar methods did not find any significant connection **[30, 31]**. This suggests that the link between LDL and breast cancer risk is still unclear and more research is needed to resolve these mixed findings.

In the context of HDL, we found no association between HDL-C levels and breast cancers, which is consistent with the findings of other studies **[31, 61, 70, 88]**. In particular, Orho-Melander, using MR, did not observe any effect of HDL-C on the risk of breast cancer in women of European ethnicity **[31]**. However, other meta-analyses and prospective studies have reported a linear association between HDL-C and breast cancer risk **[33, 30, 57, 58, 89–91]**. Additionally, some research suggests that serum HDL-C may protect against breast carcinogenesis, particularly among postmenopausal women **[24, 59, 70]**. Our findings differ from these earlier studies, which might be due to the weaker effects observed in our summary data, as this could lead to reduced statistical power. Another factor may be our inability to account for not only menopausal status but also weight and BMI which are known to interact significantly with cholesterol levels and risk of breast cancer **[92, 93]**.

## Strength and Limitations

This study possesses several significant strengths. First, it is the first study to use MR to determine the genetic association between lipids and breast cancer in African women. We included the largest GWAS to date for breast cancer among African women, consisting of 18,034 cases and 22,104 controls. The four lipid traits obtained from the AWI-GEN study were associated with strong IVs, as indicated by F-statistics well above the conventional threshold of 10 which satisfies the first assumption for MR analysis. Additionally, the population for both the exposure and outcome datasets consists entirely of African individuals, reducing potential biases related to population stratification. Using IVW, WM, MR-Egger, and Cochran Q tests, we tried our best to control pleiotropic effects, a major concern in MR studies. The significant negative association between triglycerides and overall breast cancer observed in the IVW model was consistent between the WM and MR-Egger models and the MPRESSO algorithms. These findings suggest that elevated TG levels may potentially offer a protective effect against breast cancer in African women. As we have highlighted the strengths, there are also limitations to our study. The sample size, particularly for the lipid traits, was relatively small, which may have reduced the statistical power to detect associations. Moreover, due to the limited number of GWAS studies conducted among African populations, we were unable to account for potential confounders such as BMI, obesity, and other factors known to influence breast cancer pathogenesis. Additionally, the study focused exclusively on data from individuals of African ancestry, which could limit the generalizability of the findings to other populations. Finally, more research is needed to explore the mechanisms by which elevated triglyceride levels may reduce the risk of overall breast cancer among individuals of African descent.

## Conclusions

Our study revealed that genetically elevated triglyceride levels were associated with a reduced risk of overall breast cancer in African women. Given the mixed findings in the existing literature and the unique characteristics of our study population, further research is essential to better understand the complex relationships between lipid profiles and breast cancer risk.

## Data Availability

The analysis code in R is available on request and all data displayed in figures are available in supplementary Tables 1-12.

