## Supplemental figures for "Associations Between Lipid Traits and Breast Cancer Risk: A Mendelian Randomization Study in African Women"

**Supplementary Figure 1**. Scatter plots showing the correlation of genetic associations of lipid traits with genetic associations with overall breast cancers. Each dot represents one SNP used as the genetic instrument. Coloured lines represent the slopes of the different regression analyses. Abbreviations: SNP, single nucleotide polymorphisms.

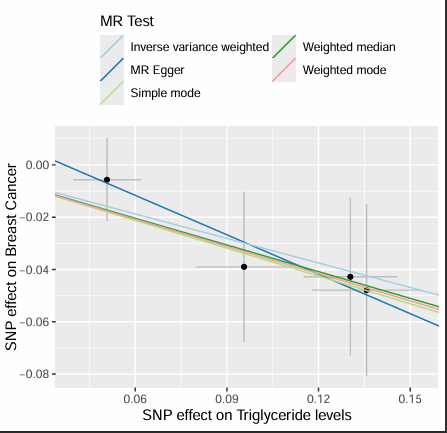

Total cholesterol

Triglyceride

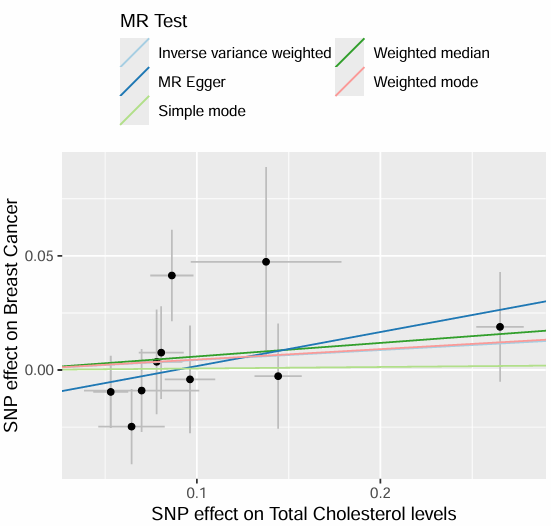

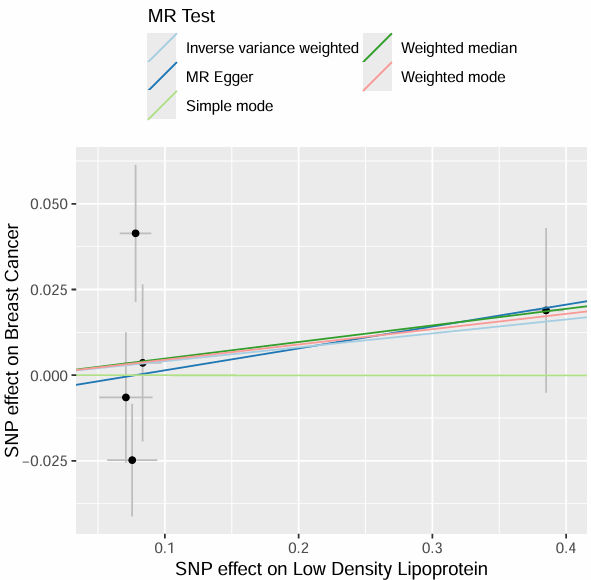

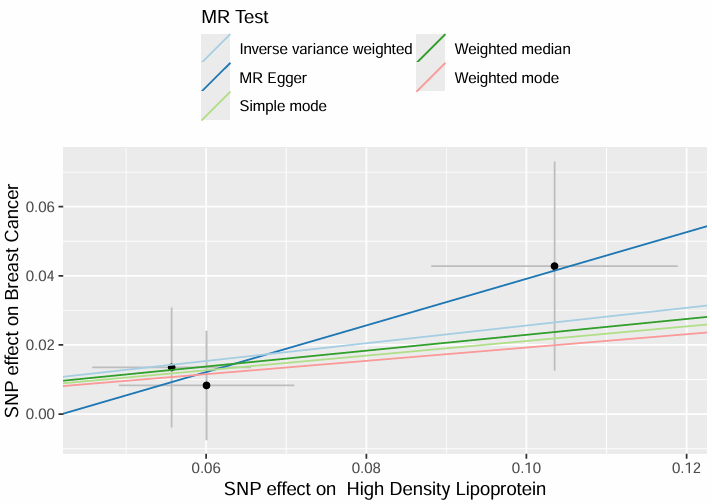

Low density lipoprotein

High density lipoprotein

**Supplementary Figure 2**. Scatter plots showing the correlation of genetic associations of lipid traits with genetic associations with estrogen positive breast cancer. Each dot represents one SNP used as the genetic instrument. Coloured lines represent the slopes of the different regression analyses. Abbreviations: SNP, single nucleotide polymorphisms; ER+, estrogen positive.

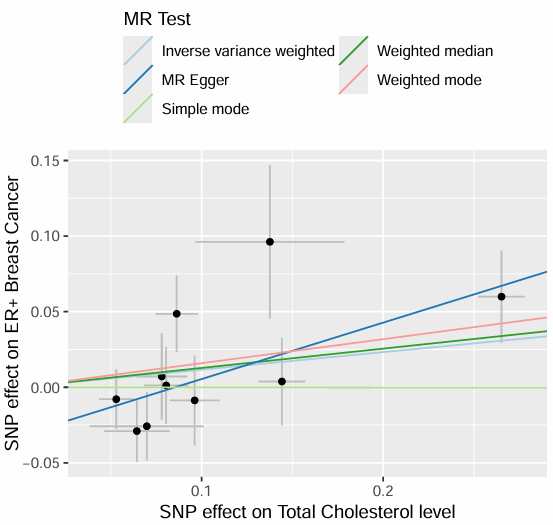

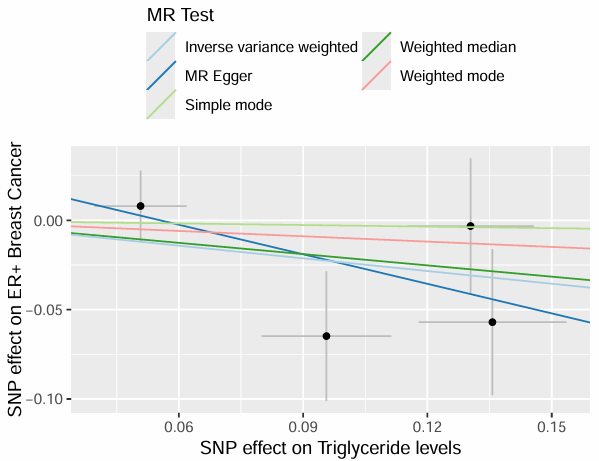

Total cholesterol

Triglyceride

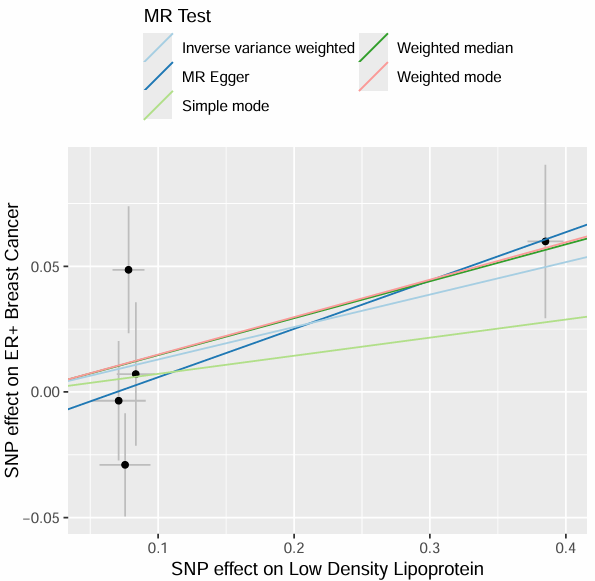

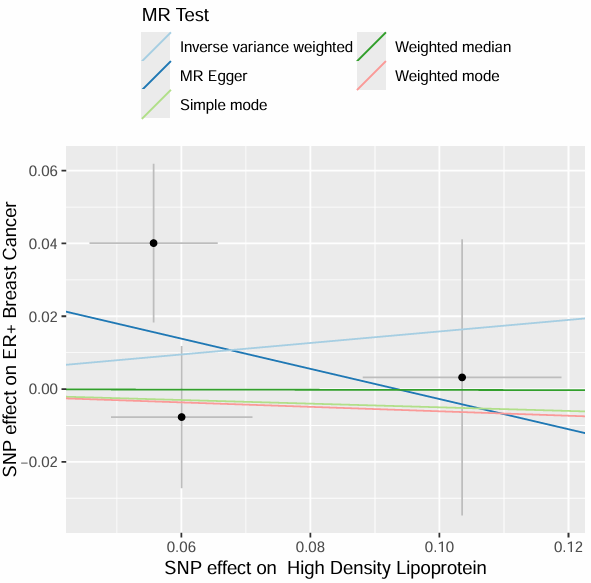

Low density lipoprotein

High density lipoprotein

**Supplementary Figure 3**. Scatter plots showing the correlation of genetic associations of lipid traits with genetic associations with estrogen negative breast cancer. Each dot represents one SNP used as the genetic instrument. Coloured lines represent the slopes of the different regression analyses. Abbreviations: SNP, single nucleotide polymorphisms; ER-, estrogen negative.

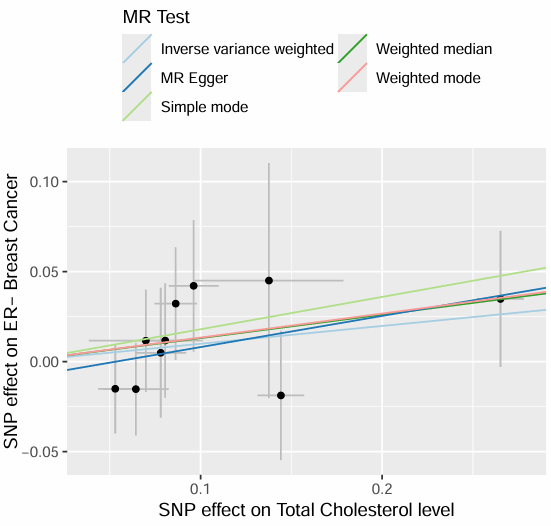

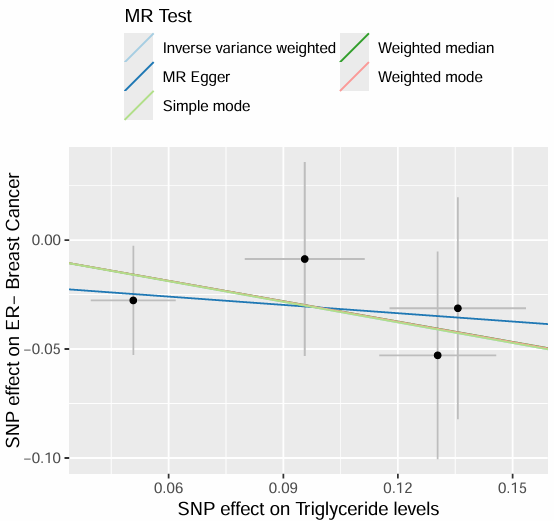

Total cholesterol

Triglyceride

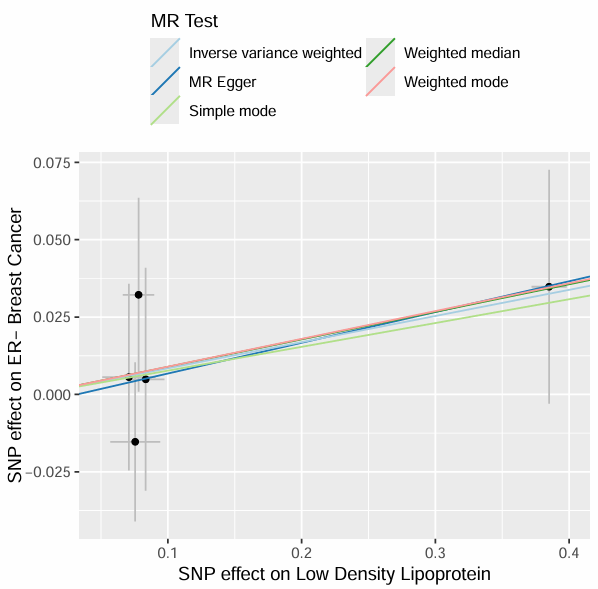

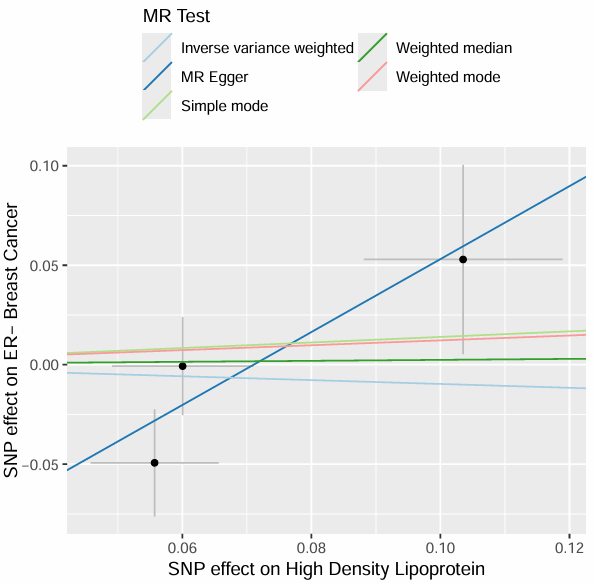

Low density lipoprotein

High density lipoprotein

**Supplementary Figure 4**. Scatter plots showing the correlation of genetic associations of lipid traits with genetic associations with triple negative breast cancer. Each dot represents one SNP used as the genetic instrument. Coloured lines represent the slopes of the different regression analyses. Abbreviations: SNP, single nucleotide polymorphisms; TNBC, triple negative breast cancer.

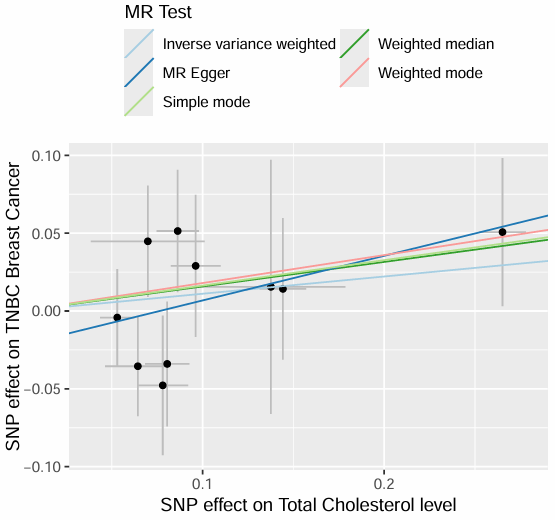

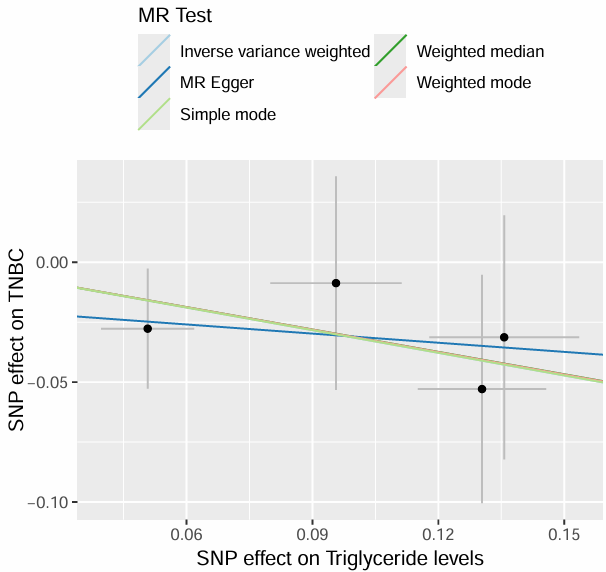

Triglyceride

Total cholesterol

Low density lipoprotein

High density lipoprotein

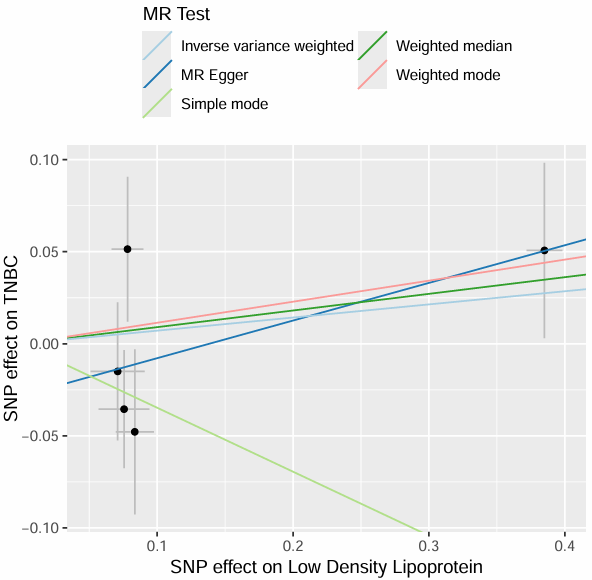

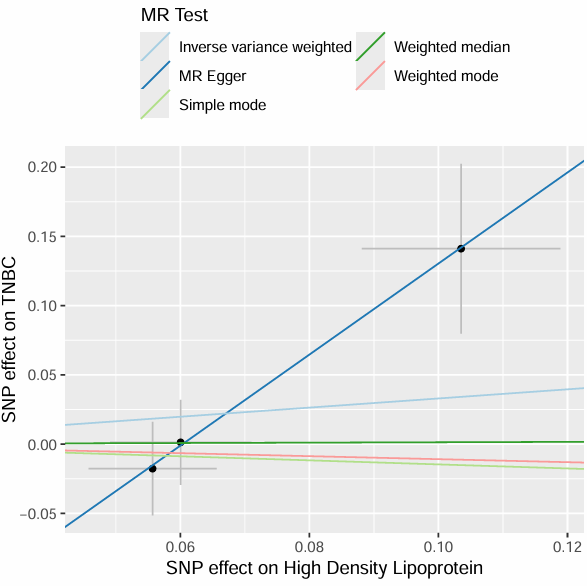

**Supplementary Figure 5**. Leave-One-Out sensitivity plots showing the impact of each SNP on the causal estimate between lipid traits and overall breast cancer. Coloured red line represents the overall causal estimate, while the black dots indicate the estimate when each SNP is individually removed.

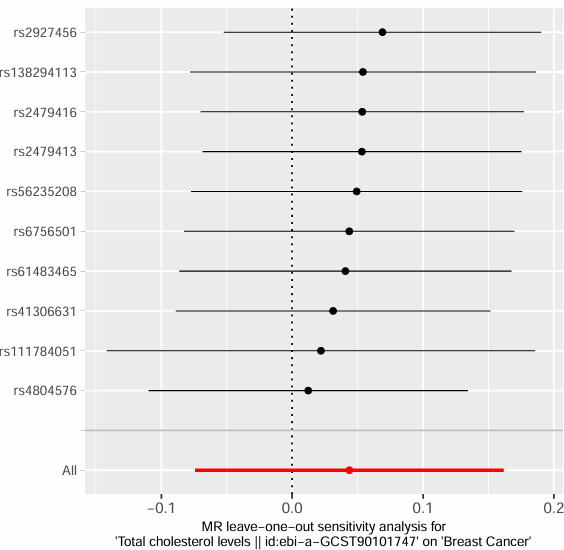

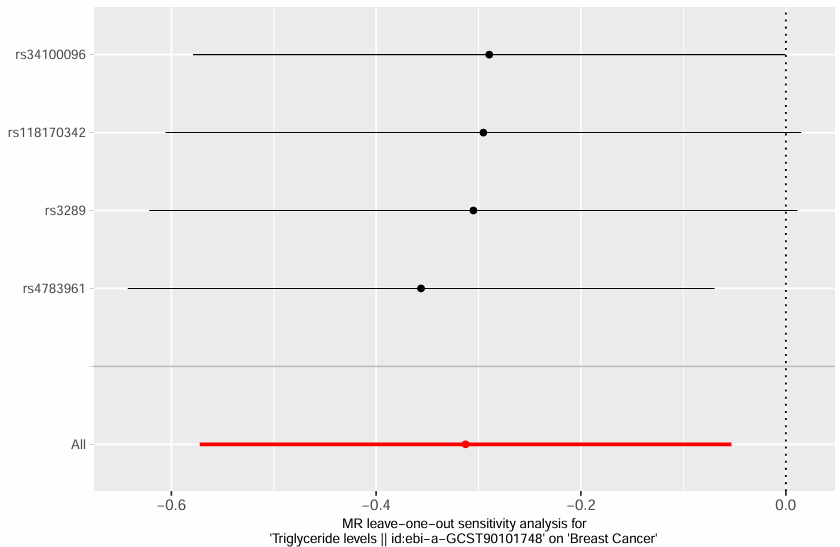

Total cholesterol

Triglyceride

Low density lipoprotein

High density lipoprotein

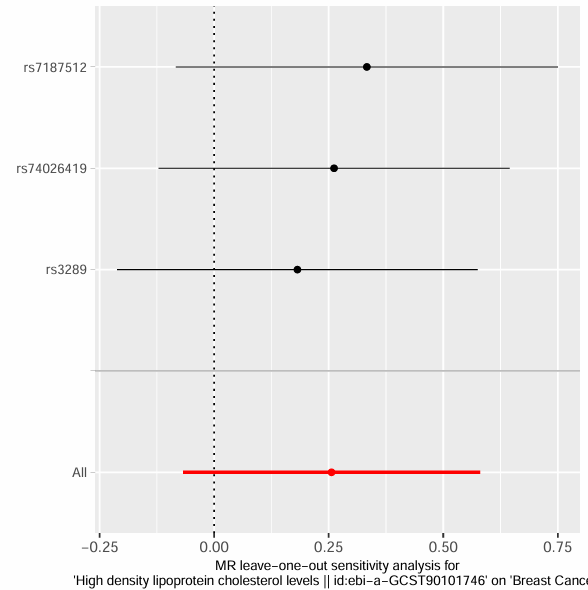

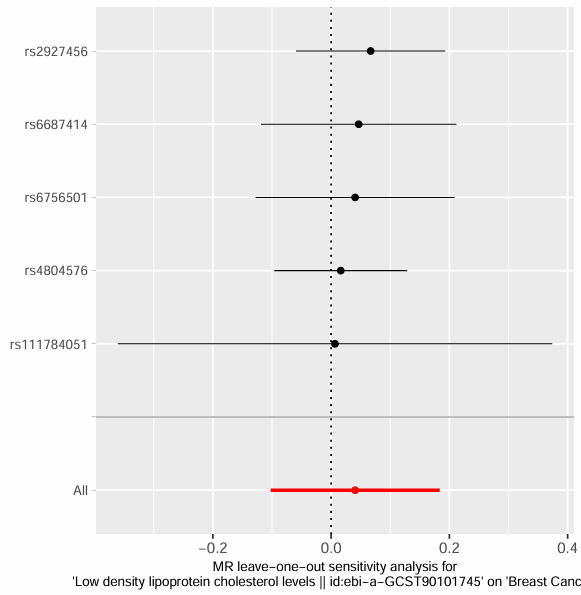

**Supplementary Figure 6**. Leave-One-Out sensitivity plots showing the impact of each SNP on the causal estimate beween lipid traits and estrogen positive breast cancer. Coloured red line represents the overall causal estimate, while the black dots indicate the estimate when each SNP is removed.

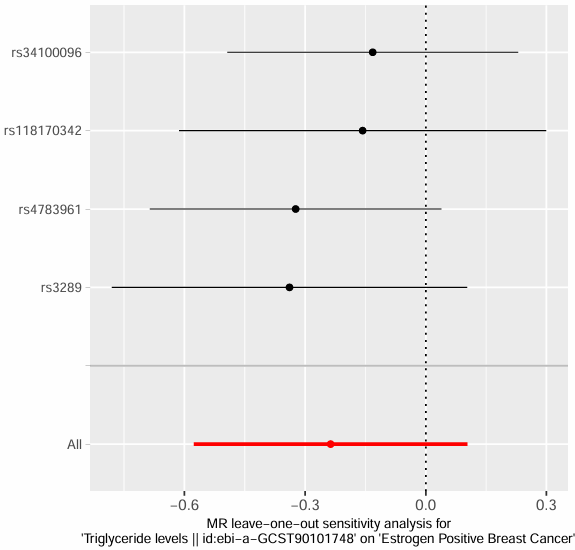

Total cholesterol

Triglyceride

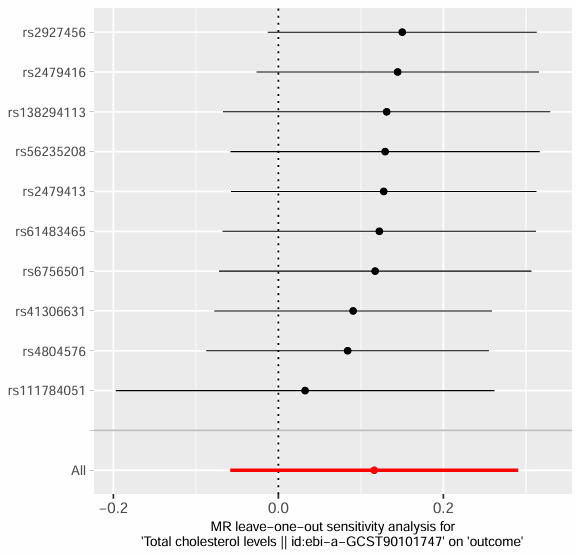

Low density lipoprotein

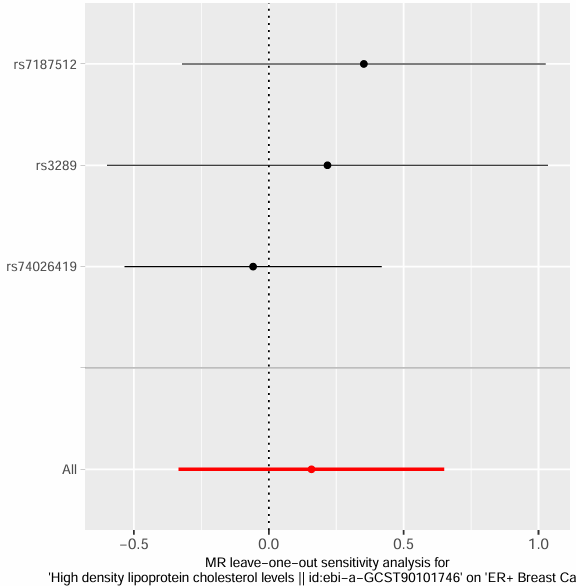

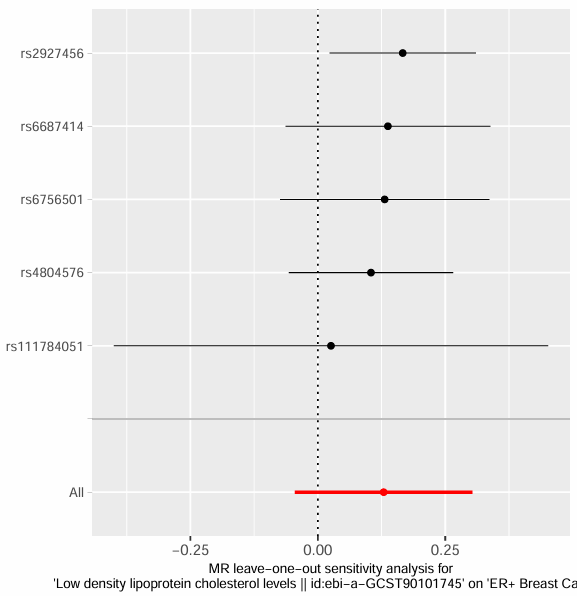

High density lipoprotein

**Supplementary Figure 7**. Leave-One-Out sensitivity plots showing the impact of each SNP on the causal estimate between lipid traits and estrogen negative breast cancer. Coloured red line represents the overall causal estimate, while the black dots indicate the estimate when each SNP is removed.

Total cholesterol

Triglyceride

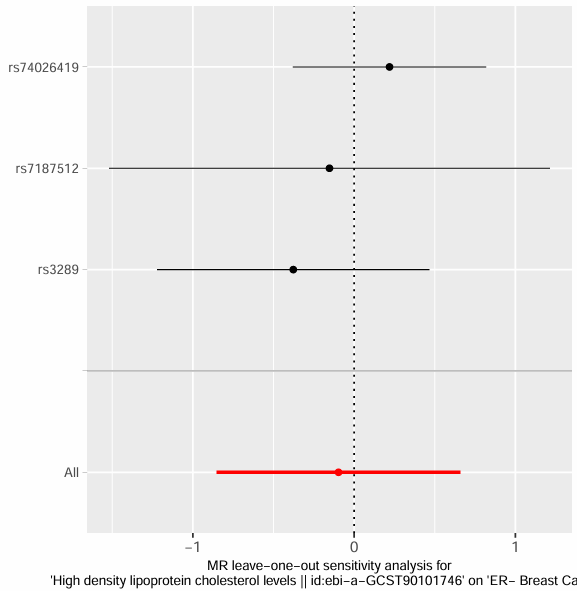

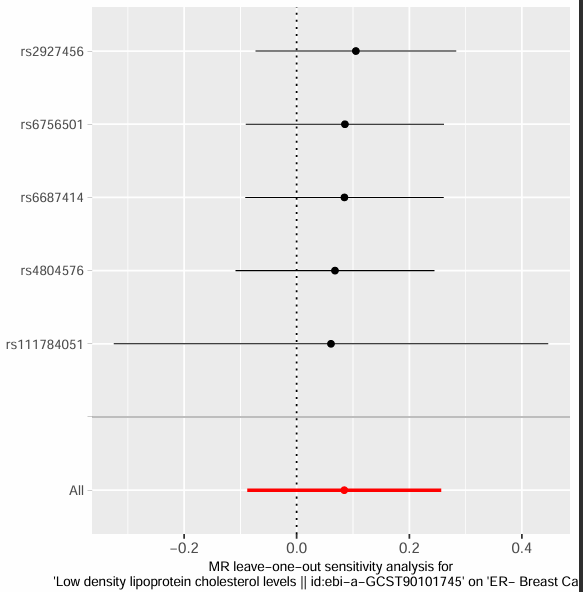

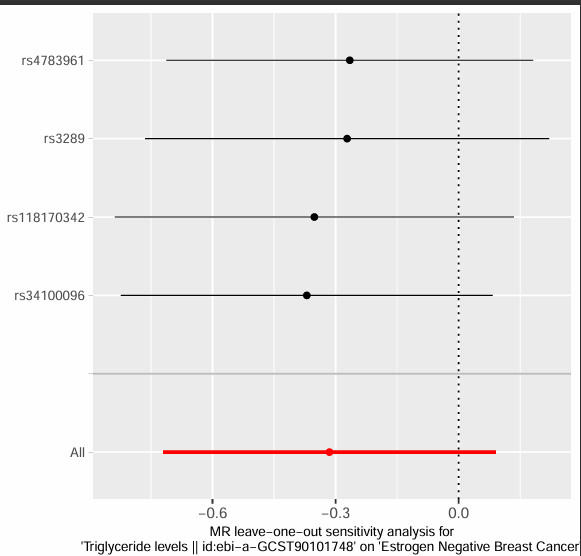

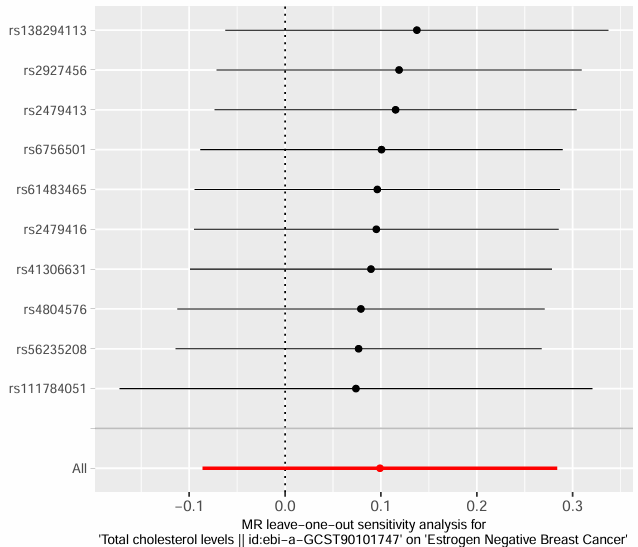

Low density lipoprotein

High density lipoprotein

**Supplementary Figure 8**. Leave-One-Out sensitivity plots showing the impact of each SNP on the causal estimate between lipid traits and triple negative breast cancer. Coloured red line represents the overall causal estimate, while the black dots indicate the estimate when each SNP is removed.

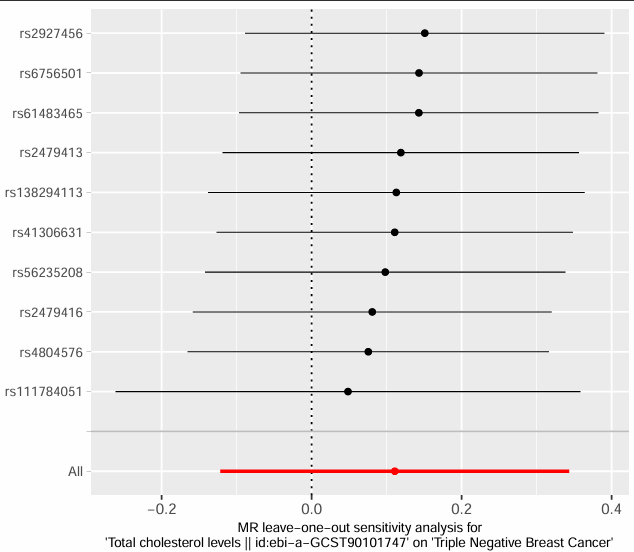

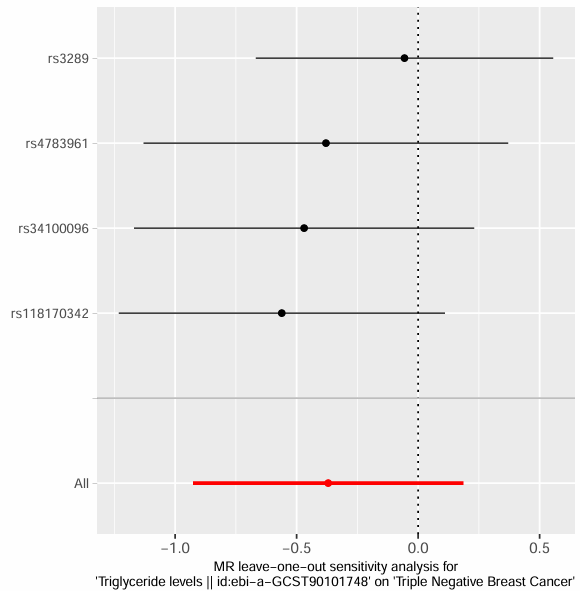

Total cholesterol

Triglyceride

Low density lipoprotein

High density lipoprotein

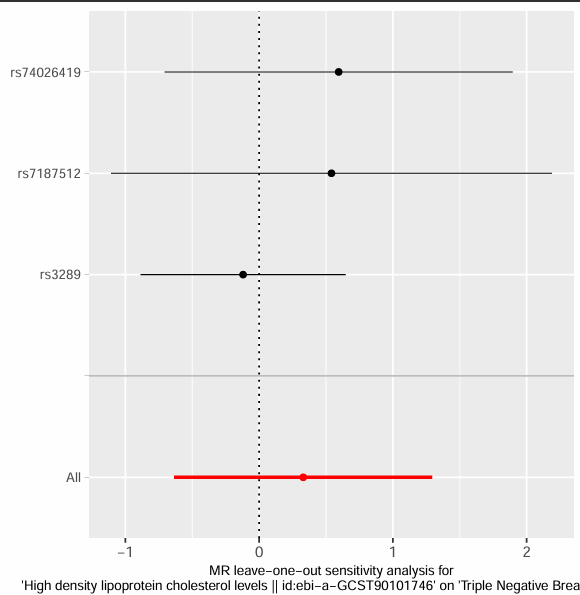

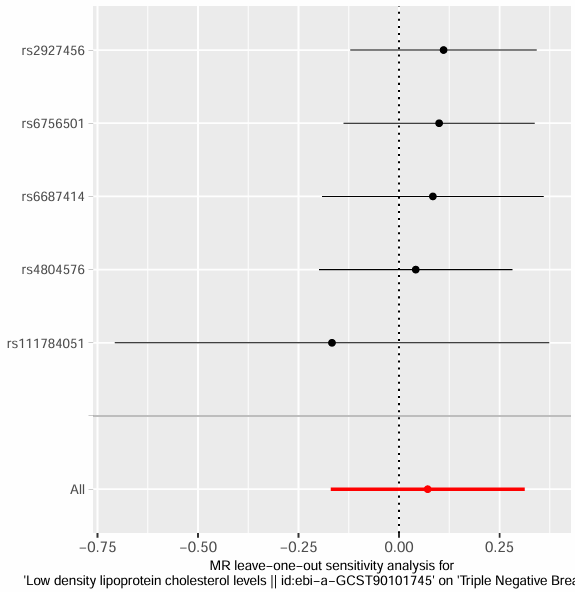
